# Skin-related adverse events and their determinants among Diabetic patients on insulin therapy in Tanzania

**DOI:** 10.1101/2025.02.25.25322829

**Authors:** Dennis P. Mbwambo, Wigilya Mikomangwa, Rajabu Hussein Mnkugwe, Bertha Mallya, Manase Kilonzi, Method Kazaura, Magreth Angelus, Kaushik Ramaiya, Mary Mayige, Ritah Mutagonda, Alphonce Ignace Marealle

## Abstract

**Background:** Globally, more than 530 million people live with diabetes mellitus (DM), and the burden continues to rise. Insulin remains the cornerstone of DM management worldwide. However, literature indicates that insulin users are at risk of developing abscesses and scar formation at injection sites. These complications may compromise adherence to therapy, thereby affecting the intended therapeutic outcomes. This study aimed to assess the prevalence of abscesses and scar formation at injection sites and their determinants among diabetic patients on insulin therapy in Dar es Salaam, Tanzania.

**Methods:** A hospital-based analytical cross-sectional study was conducted between February and May 2024. A total of 428 patients with DM on insulin therapy were enrolled from four selected hospitals in Dar es Salaam. A validated case report form (CRF) was used to collect socio-demographic characteristics and clinical data related to abscess and scar formation following insulin therapy. Data were analyzed using Stata version 15.0 software, with findings summarized as frequencies and percentages. Determinants of abscess and scar formation were assessed using modified Poisson regression, and a p-value of <0.05 was considered statistically significant.

**Results:** Out of 428 participants, 191 (44.6%) were aged over 45 years, 233 (54.4%) were female, and 185 (43.2%) had a primary education level. The prevalence of abscesses and scar formation was 95 (22.2%) and 200 (46.7%), respectively. Determinants of abscess formation included improper injection technique (aPR = 1.11; 95% CI: 1.02–1.21, p = 0.009), improper injection site rotation (aPR = 2.7; 95% CI: 1.13–6.45, p = 0.025), and the use of an insulin pen (aPR = 0.13; 95% CI: 0.04–0.48, p = 0.002). For scar formation, determinants included improper injection site rotation (aPR = 1.63; 95% CI: 1.03–2.32, p = 0.037), uncontrolled blood glucose levels (aPR = 1.69; 95% CI: 1.01–2.84, p = 0.049), and the use of insulin and syringes obtained from community drug outlets (aPR = 2.51; 95% CI: 1.22–1.98, p = 0.035).

**Conclusion:** The study identified a significant burden of abscess and scar formation among DM patients on insulin therapy. Key determinants included improper injection techniques, inadequate rotation of injection sites, and uncontrolled blood glucose levels. Regular training for DM patients on proper insulin injection practices is essential to minimize these complications.

## Introduction

Globally, the prevalence and incidence of diabetes mellitus (DM) have increased at an alarming rate (1). In 2019, it was estimated that 530 million individuals globally were living with DM. During the same period, the prevalence of DM among adults aged 20 years and above in Africa was 7.1% (2). Tanzania is ranked among the top five countries in Africa with a high prevalence of DM; currently estimated to be around 12% (2). In West Africa and Asia, the prevalence of DM among the general population is higher in urban compared to rural; the difference is estimated to be > 10% (3,4). The prevalence of DM among the general population in Dar es Salaam has increased from < 2% to almost 10% just in two decades (5).

Insulin therapy is a cornerstone for managing both type 1 and type 2 DM. In recent years, it was observed that insulin consumption has increased proportionally with the increased prevalence of DM (1). Globally, out of 463 million DM patients, 150 to 200 million patients regularly use insulin (1). The increased population of insulin users may be associated with some safety issues including skin-related adverse events like abscesses and scars at the site of insulin injection (6,7). Abscesses and scars at the insulin injection site are cosmetically unacceptable and compromise insulin absorption and therefore failure in blood glucose regulation (8). Safety surveillance of insulin users is paramount for the detection of adverse events unidentified during the registration of insulin and its accessories (9). Early detection and reporting of adverse events is crucial in mitigation of the occurrence of adverse events preventing further damage to large populations (2).

Studies from high- and middle-income countries indicate a high prevalence of skin-related adverse events among insulin users, associated with factors such as poor injection techniques, inadequate hygiene, needle length, and improper injection site rotation (10–12). However, information on skin-related adverse events, particularly abscess and scar formation among insulin users, is scarce in low- and middle-income countries (LMICs) like Tanzania, despite the country being ranked among the top five African nations with a high burden of DM patients. This study aimed to assess the prevalence of abscesses and scar formation at injection sites and their determinants among diabetic patients on insulin therapy in selected hospitals in Dar es Salaam, Tanzania.

## Materials and Methods

### Study design and settings

A hospital-based analytical cross-section study was conducted between February and May 2024 in selected hospitals in Dar es Salaam, Tanzania. Dar es Salaam region is Tanzania’s largest and most populated city [Available at https://sensa.nbs.go.tz/publication/volume1a.pdf. Accessed on 15 September 2023)]. Also, literature shows that the prevalence of DM among the general population in Dar es Salaam has increased significantly compared to other regions in Tanzania (13).

Four hospitals in Dar es Salaam with diabetic clinics were selected randomly out of 10 public and 10 private hospitals with high magnitude of DM patients; two public hospitals (Mwananyamala and Temeke Regional Referral Hospitals) and two private hospitals (Shree Hindu Mandal and Hubert Kairuki Memorial Hospital). The four hospitals are in the secondary-level category based on healthcare system of Tanzania and each facility has healthcare professionals specializing in DM. Each facility handles a significant number of DM patients monthly; At least 1007 patients at Shreel Hindu Mandal, 670 for Mwananyamala RRH, 560 for Temeke RRH and 502 for Hubert Kairuki Memorial Hospital.

### Study population

This study included patients with diabetes mellitus (Type I and Type II) who were on regular insulin therapy and attended clinics at four selected healthcare facilities. Only patients who had been using insulin for at least four weeks prior to data collection were enrolled. Patients who had pre-existing abscesses or scars at insulin injection sites before initiating insulin therapy were excluded.

### Sample size and sampling technique

A total of 428 DM patients on insulin therapy were enrolled in this study. Formula for cross-sectional study (n = p(1-p) z^2^ /d^2^) was used to calculate the sample size (14). Due to lack of similar study in Tanzania, we assumed a prevalence of 50%, a confidence level of 95% (z = 1.96), and precision (d) of 5%. A minimum of 385 participants was obtained and assuming 10% non-respondent rate a total of 428 participants was obtained.

Three-stage sampling strategy was used to obtain the study participants. Firstly, we obtained a list of the top ten (10) public healthcare facilities with a high number of DM patients served in a month for the past three months. Secondly, we randomly selected two (2) public healthcare facilities from that top ten list using a ballot box. A similar procedure was employed to obtain the two (2) private healthcare facilities from the list of top ten (10) private hospitals. Thirdly, a systematic sampling technique was employed during the recruitment of participants using a predetermined sampling interval. We recruited one eligible subject after skipping another until each facility’s sample size was attained. The number of study participants contributed by each facility depended on the ratio contributed by the facility to the total DM patients served in all four facilities.

### Data collection procedures

A case report form (CRF) was developed based on a literature review, consultations with experts, and the investigators’ experience in conducting quantitative research. The CRF was used to collect data from patients’ files and during follow-up, including sociodemographic characteristics and information related to abscess and scar occurrence. This included details on skin allergies, abscess and scar occurrences, the source of insulin and syringes, needle length, reuse of needles, insulin storage, average blood glucose levels, and the presence of comorbidities.

The tool was pretested with 30 participants at Amana Regional Referral Hospital (RRH), which was not one of the study sites but is a secondary healthcare facility in Dar es Salaam with similar settings to the study sites. The pretest helped refine and modify the tool before the start of data collection.

Four research assistants (one per site) were recruited and trained to collect data using the CRF. These research assistants held degrees or diplomas in medicine, or nursing and worked in the diabetic clinics of their respective study sites. To enhance their understanding of the study and familiarize them with the tool, the research assistants participated in the pretesting phase.

Data collection was performed on daily basis during the regular patient clinic visits. Before enrollment, eligible patients were sensitized about the study. Those who showed interest were given the consent form to read and understand. Both English and Swahili versions were used at the subject’s convenience. Those incapables of reading were supported by their guardian or the study team. Following an understanding of the study, patients who were willing to join the study were asked to sign or thumb the consent form. Using the CRF, the research assistant obtained the required information from the patient file and a physical examination was performed to observe the abscess and scar at the insulin injection site. Patients were asked to provide information on whether they developed abscesses or scars in the past 12 months from the date of the interview. Physical examinations of the adverse event were held in a private room to ensure the client’s comfort and confidentiality. Furthermore, each participant was assigned a unique identification number with a two-letter facility prefix, e.g., TM001. The interview could take an average of 6-12 minutes per client. Three-month random blood glucose readings were taken from the patient’s files available at the clinic. The readings were used to estimate the patient’s average blood glucose.

Assessment of patients’ awareness of body parts recommended for injecting insulin was done by asking them to mention all such body parts. Those who managed to mention all the recommended parts: abdomen, thigh, upper hand, and buttocks were considered to be aware, and those who failed to mention all the parts were considered unaware. The correctness of injection site rotation was assessed by considering if the patient divided the abdomen into four quadrants, utilizing each quadrant for a week before shifting to another quadrant. The other remaining body parts need to be divided into two halves and each half to be injected for one week. Additionally, the two injection sites should be at least 1 cm apart.

### Data analysis

Data were entered into MS Excel and transferred to Stata version 15.0 for analysis. Frequency and percentages were used to summarize the findings. While determinants of abscess and scar formation were assessed systematically using the chi-square test and those variables with the p-value of < 0.2 were taken into multivariate modified Poisson regression model and variables with a p-value < 0.05 were considered determinants.

### Ethics statement

The study protocol was reviewed and approved by the Muhimbili University of Health and Allied Sciences (MUHAS)-Research and Ethics Committee (MUHAS-REC) with a reference number DA282/298/01.C/2057 The medical officer in charge of the respective health facilities permitted to collect data. Each participant gave informed consent before data collection and ethics were observed through data collection as stipulated in the Declaration of Helsinki.

## Results

### Description of the study participants

A total of 428 diabetic patients were enrolled in the study. The mean age of the participants was 38.4 years (SD ±19.4), with the majority being adults aged ≥45 years (44.6%). Females comprised 54.4% (233) of the participants, 59.6% (255), were unemployed, and 70.3% (301) had health insurance coverage. Additionally, 1.9% of the participants were smokers, and 6.3% reported alcohol consumption, (table 1).

**Table 1:** Sociodemographic and clinical characteristics of study participants.

| Variables | Category | Frequency | Percent |
| --- | --- | --- | --- |
| Age (years) | ≤ 24 | 141 | 33.0 |
|  | 25-44 | 96 | 22.4 |
| | ≥ 45 | 191 | 44.6 |
| Sex | Male | 195 | 45.6 |
|  | Female | 233 | 54.4 |
| Level of education | Non-primary education | 185 | 43.2 |
|  | Secondary education | 182 | 42.5 |
|  | University | 61 | 14.3 |
| Patient occupation | Unemployed | 255 | 59.6 |
|  | Self-employed | 109 | 25.5 |
|  | Employed | 64 | 14.9 |
| Health insurance | Have health insurance | 301 | 70.3 |
|  | No health insurance | 127 | 29.7 |
| Cigarette smoking | Yes | 8 | 1.9 |
|  | No | 420 | 98.1 |
| Alcohol consumption | Yes | 27 | 6.3 |
|  | No | 401 | 93.7 |

### Skin-related adverse events and related information among DM patients on insulin injection

Of the 428 study participants, the prevalence of abscess formation was 95(22.2%) and 200(46.7%) for scar occurrence. The majority, 378(88.3%) of participants, obtained their insulin and syringes from the facility, and 398(93%) reused the syringes. Most 52(54.7%) of the observed abscesses are mild and the majority 207(48.4%), occur following the use of mixed/fused insulin (table 2).

**Table 2:** Prevalence of abscess and scar at injection site among DM patients on insulin therapy (n =428).

| Variables | Category | Frequency | Percent |
| --- | --- | --- | --- |
| Skin allergy | Yes | 47 | 11.0 |
|  | No | 381 | 89 |
| Ever developed abscess | Yes | 95 | 22.2 |
|  | No | 333 | 77.8 |
| When abscesses happened (n=95) | Between Jan-Mar 2023 | 33 | 34.7 |
|  | Between April-Jun 2023 | 12 | 12.6 |
|  | Between July-Sept 2023 | 16 | 16.8 |
|  | Between Oct-Dec 2023 | 34 | 35.9 |
| Seriousness of the abscess (n=95) | Mild resolved within a few days | 52 | 54.7 |
|  | Needed Medication | 36 | 37.9 |
|  | Severe Needed Hospitalization | 7 | 7.4 |
| Ever developed scar at the injection site | Yes | 200 | 46.7 |
|  | No | 228 | 53.3 |
| When scars happened (n=200) | Between Jan-Mar 2023 | 45 | 22.5 |
|  | Between April-Jun 2023 | 23 | 11.5 |
|  | Between July-Sept 2023 | 24 | 12 |
|  | Between Oct-Dec 2023 | 108 | 54 |

### Patient awareness of insulin injection procedures and their injecting practices

Twenty-three percent (102/428) reported having other comorbidities other than DM. Of the study participants, 11% (47) reported having skin allergy. The most common syringe used for insulin injection had a needle length of ≤ 6mm accounting for 55.6%. The majority of participants, 93% (398/428), reported reusing syringe needles. The reuse frequency per a single needle varied from one shot to more than three shots per needle. The majority of the study participants, 88% (378/428) obtained the insulin and syringes at the health facilities they attended clinic. The majority of the study participants, 79.4% (339/428) adhered to the insulin storage conditions. At least 65% (278/428) of study participants know all the body parts recommended for optimum insulin absorption and 90.2% (386/428) of the study participants demonstrated the ability to inject insulin correctly. However, 63.2% (273/428) of the study participants did not know how to perform injection site rotation. Of the study participants, 32.2% (138/428) always cleaned the injection site before injecting the insulin. The study also found that 79.4% (340/428) of study participants had poorly controlled average blood glucose (Table 3).

**Table 3:** Patient awareness of insulin injection procedures and their injecting practices.

| Variables | Category | Frequency | Percent |
| --- | --- | --- | --- |
| The source of insulin and syringes | At the health facility | 378 | 88.3 |
|  | Other Places | 50 | 11.7 |
| Type/insulin formulation | Single separate formulations | 95 | 22.2 |
|  | Mixed/fused Formulation | 207 | 48.4 |
|  | Insulin Pen | 126 | 29.4 |
| Needle length in 'mm" (n=378) | $\leq 6$mm | 238 | 55.6 |
|  | Insulin pen needle | 125 | 29.2 |
|  | Unknown | 65 | 15.2 |
| Ever reused syringe needle | Yes | 398 | 93 |
|  | No | 30 | 7 |
| How often reused the needle (n=398) | Sometimes | 134 | 31.3 |
|  | Often | 137 | 32 |
|  | Always | 127 | 29.7 |
| Frequency of needle re-use<br>(n=398) | Up to three shorts | 217 | 57.7 |
|  | More than three shorts | 181 | 42.3 |
| Aware of injection sites | Know all the sites | 278 | 65 |
|  | Know a few of the sites | 150 | 35 |
| Insulin injecting technique | Correct | 386 | 90.2 |
|  | Not correct | 42 | 9.8 |
| Injection site rotation | Rotates correctly | 155 | 36.2 |
|  | Does not rotate correctly | 273 | 63.8 |
| Clean injection site site | Sometimes | 290 | 67.8 |
|  | Always | 138 | 32.2 |
| Cleaning procedure (n=253) | Correct | 165 | 65.2 |
|  | Not correct | 88 | 34.8 |
| Average blood glucose | Well-controlled | 88 | 20.6 |
|  | Not controlled | 340 | 79.4 |
| Insulin storage | 2 to 8°C | 339 | 79.4 |
|  | >8°C | 89 | 20.6 |
| Presence of any comorbidities | Yes | 102 | 23.8 |
|  | No | 326 | 76.2 |

### Factors associated with abscess formation at the insulin injection site among DM patients

Those who were self-employed had a 134% higher prevalence than the non-employed (aPR = 2.34; 95%CI: 1.04-5.27; p = 0.4). Patients reported with improper injection rotation had a 170% higher prevalence compared to those rotated the injection sites (aPR = 2.7; 95%CI: 1.13-6.45; p = 0.025). Those reported with improper injection technique had an 11% higher prevalence compared to those who adhered to proper injection technique (aPR=1.11; 95%CI: 1.02-1.21; p = 0.009). Those who used insulin pens had 87% lower prevalence when compared to those who used vials and syringes (aPR = 0.13; 95%CI: 0.04-0.48; p = 0.002) (Table 4).

**Table 4:** Factors associated with abscess formation at insulin injection site.

| Variable | N | Abscess<br>formation (%) | Unadjusted |  | Adjusted |  |
| --- | --- | --- | --- | --- | --- | --- |
|  |  |  | cPR,95% CI | p-value | aPR,95% CI | p-value |
| Age group |  |  |  |  |  |  |
| ≤24 | 141 | 42 (29.79) | Ref |  |  |  |
| 25-44 | 96 | 24 (25.00) | 0.83(0.96-1.09) | 0.41 | 0.99 (0.65-1.53) | 0.99 |
| ≥45 | 191 | 29 (15.18) | 0.51(1.03-1.14) | 0.002 | 0.60 (0.99-1.15) | 0.06 |
| Sex |  |  |  |  |  |  |
| male | 195 | 45 (23.08) | Ref |  |  |  |
| female | 233 | 50 (21.46) | 0.93(0.97-1.06) | 0.689 | 1.02(0.97-1.08) | 0.87 |
| Occupation |  |  |  |  |  |  |
| Non | 255 | 51 (20.00) | Ref |  |  |  |
| Self-employed | 109 | 30 (27.5) | 1.38(0.93-2.04) | 0.290 | 2.34 (1.04-5.27) | 0.04 |
| Employed | 64 | 14 (21.88) | 1.09(0.64-1.84) | 0.572 | 1.14 (0.39-3.32) | 0.815 |
| Source of insulin & syringes |  |  |  |  |  |  |
| Health facility | 378 | 79 (20.90) | 0.94(0.86-1.02) |  | 0.91 (0.79-1.03) | 0.13 |
| Other sources | 50 | 16 (32.00) | Ref | 0.119 |  |  |
| Site rotation |  |  |  |  |  |  |
| Yes | 155 | 28 (18.06) | Ref |  |  |  |
| No | 273 | 67 (24.54) | 1.36(0.91-2.01) | 0.108 | 2.7 (1.13-6.45) | 0.025 |
| Insulin formulation |  |  |  |  |  |  |
| Single vial | 95 | 18 (18.95) | Ref |  |  |  |
| Mixed/fused | 207 | 66 (31.88) | 1.68(1.06-2.67) | 0.034 | 1.45 (0.59-3.53) | 0.42 |
| Insulin Pen | 126 | 11 (8.73) | 0.19(0.23-0.93) | 0.012 | 0.13 (0.04-0.48) | 0.002 |
| Needle length |  |  |  |  |  |  |
| ≤ 6mm | 238 | 69 (28.99) | 3.30 (1.01-1.05) | 0.32 | 0.94 (0.83-1.08) | 0.43 |
| Unknown | 65 | 15 (23.08%) | 2.60 (0.85-0.98) | <0.01 | 1.12 (0.97-1.29) | 0.11 |
| Pen needle | 125 | 11 (8.80) | Ref |  |  |  |
| Injecting technique |  |  |  |  |  |  |
| Correct | 386 | 178 (46.11) | Ref |  |  |  |
| Not correct | 42 | 22 (52.38) | 1.11 (1.07-1.16) | <0.01 | 1.11 (1.02-1.21) | 0.009 |

### Factors associated with scar formation at the insulin injection site among DM patients

Those who did not perform proper injection site rotation had 63% higher prevalence than those who adhered to proper injection site rotation (aPR = 1.63; 95%CI: 1.03-2.32; p = 0.037). Patients with poorly controlled blood glucose had a 69% higher prevalence compared to those with controlled blood glucose (aPR = 1.69; 95%CI:1.01-2.84, p = 0.049). Participants who were self-employed had a 100% higher prevalence compared to the unemployed (aPR = 2.0; 95%CI:1.21-3.33, p= 0.007). We also noted that DM patients who consumed insulin and the accessories outsourced from other places other than at the health facility they attended the clinic (community drug outlets), had 151% higher prevalence when compared to those who consumed insulin and accessories obtained at the health facilities, they attended clinics (aPR = 2.51; 95%CI: 1.22-1.98, p= 0.035) (Table 5).

**Table 5:** Factors associated with scar formation at insulin injection site.

| Variable | Total | Developed scar (%) | Crude estimates |  | Adjusted estimates |  |
| --- | --- | --- | --- | --- | --- | --- |
|  |  |  | cPR,95% CI | p-value | aPR,95% CI | P-Value |
| Age |  |  |  |  |  |  |
| ≤24 | 141 | 68 (48.23) | Ref |  |  |  |
| 25-44 | 96 | 50 (52.08) | 1.07 (0.89-1.06) | 0.561 | 0.96 (0.88-1.05) | 0.44 |
| ≥45 | 191 | 82 (42.93) | 0.89 (0.96-1.11) | 0.340 | 1.02 (0.95-1.10) | 0.40 |
| Sex |  |  |  |  |  |  |
| Male | 195 | 93 (47.69) | Ref |  |  |  |
| female | 233 | 107 (45.92) | 0.96 (0.95-1.07) | 0.715 | 0.99 (0.93-1.05) | 0.79 |
| Occupation |  |  |  |  |  |  |
| Un employed | 255 | 108 (42.35) | Ref |  |  |  |
| Self-employed | 109 | 59 (54.13) | 1.65 (1.02-2.52) | 0.360 | 2 (1.21-3.33) | 0.007 |
| Employed | 64 | 33 (50.09) | 1.45 (0.84-2.51) | 0.831 | 1.9 (1.04-3.41) | 0.036 |
| Source of insulin & syringes |  |  |  |  |  |  |
| At health facility | 378 | 166 (43.92) | Ref |  |  |  |
| Other sources | 50 | 34 (68.00) | 2.71 (1.45-5.09) | 0.001 | 2.51 (1.22-1.98) | 0.035 |
| Injection site rotation |  |  |  |  |  |  |
| Yes | 155 | 66 (42.58) | Ref |  |  |  |
| No | 273 | 134 (49.08) | 1.30 (0.87-1.93) | 0.191 | 1.63 (1.03-2.32) | 0.037 |
| **Presence of comorbidity** |  |  |  |  |  |  |
| Yes | 102 | 40 (39.22) | 1.07 (0.99-1.14) | 0.073 | 1.03 (0.96-1.11) | 0.35 |
| No | 326 | 160 (49.08) | Ref |  |  |  |
| **Insulin storage** |  |  |  |  |  |  |
| 2 to 8°C | 339 | 150 (44.25) | Ref |  |  |  |
| >8°C | 88 | 49 (55.68) | 1.26 (0.86-1.01) | 0.061 | 0.95 (0.88-1.03) | 0.21 |
| **Blood glucose level** |  |  |  |  |  |  |
| Controlled | 88 | 31 (35.23) | Ref |  |  |  |
| Uncontrolled | 340 | 169 (49.71) | 1.82 (1.12-2.96) | 0.015 | 1.69 (1.01-2.84) | 0.049 |

## Discussion

The present study assessed the prevalence of abscess and scar formation at the insulin injection site and associated factors among DM patients who attended clinics at selected private and public hospitals in Dar-es-Salaam, Tanzania. The prevalence of abscesses 22.2%, observed in this study is higher compared to what was reported from the USA and England (10,16). According to these studies, the prevalence of abscesses was reported to be rare. One of the possible explanations for the variability in prevalence could be variability in study location, study population, and the health care systems. Further, the study done in England involved only children while this study captured all the age groups. The findings from the USA reported by Richardson were obtained from the study done many years ago this could be another possible explanation for the variability in the observed prevalence of abscesses. The increase in prevalence of the adverse event could have some association with the increase of insulin consumers; insulin is currently consumed by both the DM type 1 and also by DM type 2 patients (1).

The large prevalence of scars 46.7%, observed from this study is congruent with findings from the USA (10), where Richardson, et al., reported a prevalence of 48%. This implies that the problem does exist globally irrespective of geographical location, economic status, or ethnicity. Therefore, performing pharmacovigilance to patients on long-term use of medication including insulin consumers is paramount. Pharmacovigilance will ensure early detection of adverse events and therefore prevent further damage to a large population. However, drug safety surveillance and reporting rate is low in Tanzania (17). Poor reporting can musk the magnitude of the ongoing problem allowing it to affect a large population.

We observed that failure to perform the proper injection site rotation had a 170% and 63% higher prevalence of abscesses and scars respectively. This finding corresponds to what was observed in Ethiopia (11) and India (1). Similar to what we observed in this study, Negashi, et al., and Barua, et al., reported a fair practice of injection site rotation among DM patients. This may imply the similarities in practice among insulin users across the sub-Saharan and Asia. Failure to adhere to injection site rotation leads to trauma following the repeated shorts at a single skin segment resulting in abscesses (11,12).

A positive association between abscess and improper insulin injection techniques specifically the injection angle observed in our study, (a 170% higher prevalence) supports the findings reported from Bangladesh (12), and India (1). Despite the differences in the study design, participants, and geographical locations this study, and the former two studies revealed a significant positive association between abscess and the improper insulin injection technique. It is recommended that the syringe be tilted at a suitable angle depending on the patient’s skin thickness (18), contrary to that can lead to abscess and even failure of the insulin execution.

Occupational work was positively associated with abscesses and scars a 134% and 100% higher prevalence respectively among the self-employed than non-employees. We did not find a plausible mechanism for the observed association, however, Esmail et, al., reported that some of the occupational works are associated with skin diseases (19). However, Esmail did not explore the abscess or scars more, nor did our study go further on the types of occupational work. Therefore, this association remains pending unless further studies are done to provide clear information.

An eighty-seven percent lower prevalence of abscess at insulin injection sites among DM patients using insulin pens compared to those who use conventional syringes and needles implies that insulin pens are protective. This is not a new phenomenon because similar observations have been reported in Saudi Arabia (20) and Lebanon (20). Studies suggest that insulin pens influence easy insertion, less pain, and less skin trauma therefore, it is a user friend among DM patients on regular use of insulin therapy (21,22).

We noted that DM patients who consumed insulin and syringes outsourced from places other than the health facility they attended the clinic, had a 151% higher prevalence when compared to those who consumed insulin and accessories obtained at the health facilities. This association can have different explanations; including insufficient instructions provided by the healthcare providers in community drug outlets due to lack of knowledge on DM and its management (23–26). For example, in Addis Ababa, only 15.3%(300) of the community pharmacy professionals could provide proper counseling on DM (23). Surprisingly, even those with moderate knowledge had poor practice as observed in Saudi Arabia and China (24,25). In Ethiopia, only 17.% of those reported with moderate knowledge were reported with good practice in DM counseling (26).

Failure to control and maintain the blood glucose level at the recommended range had a positive association with scar formation at the insulin injection site among DM patients. We did not get a clear scientific explanation for this occurrence however, it is not a new phenomenon (27,28). Establishing the temporal relationship between the abscess and unregulated blood sugar was also difficult, which is considered the study’s weakness.

## Conclusions

The prevalence of abscesses and scars at the site of insulin injection among diabetic patients is high. We observed that abscess was associated with the improper injection site rotation, and improper injection techniques while using an insulin pen were protective against abscess. Scars at the insulin injection site among DM patients were associated with improper injection site rotation, uncontrolled blood sugar, and the use of insulin and accessories outsourced from community drug outlets. We recommend the Ministry of Health and stakeholders to emphasize on use of insulin pens. Community drug outlet healthcare staff should be trained on DM and its management to properly counsel DM patients. Routine blood glucose monitoring should be emphasized among DM patients.

## Data Availability

All data produced in the present work are contained in the manuscript

## Acknowledgments

The authors sincerely thank the Muhimbili University of Health and Allied Sciences–Research and Ethics Committee (MUHAS-REC) for providing ethical clearance and administrations of the visited hospitals (Mwananyamala, Temeke, Shiree Hindumandal and Kairuki) for providing permission for data collection. We also thank each appointed hospital HCP for their guidance during the data collection. Additionally, we thank the Department of Clinical Pharmacy and Pharmacology at Muhimbili University of Health and Allied Sciences for their support.

## Author contributions

Conceptualization: Dennis Patson Mbwambo

Data curation: Dennis P. Mbwambo

Formal analysis: Dennis P. Mbwambo, Peter Kunambi, Alphonce Marealle

Funding acquisition: Dennis P. Mbwambo

Methodology: Dennis P.Mbwambo, Alphonce Marealle, Wigilya Mikomangwa.

Supervision: Dennis P. Mbwambo, Alphonce Marealle, Ritah Mutagonda

Project administration: Dennis P Mbwambo

Validation: Dennis P.mbwambo

Writing-original draft: Dennis P.Mbwambo, Alphonce Marealle, Wigillya Mikomangwa, Manase Kilonzi, Method Kazaura, Writing-review & editing: Dennis P. Mbwambo, Rajabu Hussein Mnkugwe, Bertha Mallya, Manase Kilonzi, Method Kazaura, Magreth Angelus, Kaushik Ramaiya, Mary Mayige & Ritah Mutagonda & Alphonce I. Marealle.

Funding: Ministry of Health of Tanzania

Competing interest: None

Data availability statement: All data are fully available without restriction

## Notes

### Competing Interest Statement

The authors have declared no competing interest.

### Funding Statement

Self-funded study

### Author Declarations

Muhimbili University of Health and Allied Sciences Institutional Review Board (MUHAS-IRB) Ref DA282/298/01.C/2057

